# Comparing the Therapeutic Effects of Cisternostomy versus Decompressive Craniectomy in the Management of Traumatic Brain Injury - Systematic Review and Meta-analysis Protocol

**DOI:** 10.1101/2023.03.06.23286840

**Authors:** Victor Meza Kyaruzi, Tumusifu Manegabe jean de Dieu, Samuel Oreoluwa David, Larrey Kasereka Kamabu, Chibuikem A. Ikwuegbuenyi, Emmanuel Mukambo, Kwadwo Ofori Darko, Tunde A. Olabatoke, Tcheutchoua Kathy Foka, Marwa S Emhemed, Tamara Tango, Murad Othman Omran, Emmanuel Oyesiji, Wesley Harrisson Bouche Djatche, Saad Javed, Emmanuel Mduma, Emnet Tesfaye Shimber, Abdul-Aziz Abdellah Hussein, Fortune Gankpe, Costansia Anselim Bureta, Nicephorus Rutabasibwa, Lemeri L Mchome, Ignatius Esene, Osama Abdelaziz

**Author notes:** **Correspondence to;** Dr Victor Meza Kyaruzi, Department of Surgery, Muhimbili University of Health and Allied Sciences, Dar es salaa, Tanzania.

## Abstract

**Background:** Traumatic brain injury (TBI) is a critical problem which portends an intensive burden with increased mortality and disability affecting more the young population worldwide. The primary goal of TBI treatment is to control intracranial pressure (ICP) and to prevent he devastating effect of secondary brain insult. For over a hundred years decompressive craniectomy has been a standard surgical intervention for treatment of TBI, however it is not without harm since it causes serious complications including meningitis, subdural hygroma, hydrocephalous and increased reoperation rate. Cisternostomy is the most recently introduced intervention for management of cerebral edema Cisternostomy has proven its efficiency a standalone treatment as an adjunctive to decompressive craniectomy in treatment of severe traumatic brain injury. This review aims at investigating the therapeutic effects of cisternostomy when used independently or as an adjunctive to decompressive craniectomy (DC) across the available randomized clinical trials (RCTs) and non randomized studies of effect intervention (NRSI) sectional studies to optimize the strength of evidence for underpinning the strategy for treatment of traumatic brain injury.

**Methods and Analysis:** We will conduct the systematic review and meta-analysis by employing the provisions of Preferred Reporting Items for Systematic Review and Meta-analysis (PRISMA) 2020 guideline and the review protocol has been submitted to International Prospective Register of Systematic Reviews (PROSPERO) for registration before commencement of the study. We will construct the search strategy using the all field terms, medical subheading terms [MeSH Terms] with all permutations combined with Boolean operators such as AND and OR. PubMed, EMBASE, Scopus, COCHRAINE, Web Of Science, Global Index Medicus, Semantic Scholar and Google Scholar electronic databases will be searched.

**Ethical Consideration and Dissemination:** This review will not include any human participant such that the ethical clearance approval is not applicable. The protocol of this review has been registered at PROSPERO ID <u>CRD42023400894</u>. We will disseminate the final report of this review to local and international scientific conferences and The results of this review will be submitted for publication in the Journal of Neurotrauma.

## INTRODUCTION

Traumatic brain injury is a critical public problem which portends an intensive burden with increased mortality and disability affecting more the young population worldwide. Traumatic brain insults manifest into two domain forms classified as primary and secondary brain injury. The severity of primary brain injury is influenced by the extent of mechanical forces and physical impact on the cranium and brain parenchyma(1,2).Secondary brain injury occurs as a complex systemic insult triggered by increased brain edema, increased intracranial pressure and neuroendocrine derangements. The primacy goal of TBI treatment is to control ICP and to prevent he devastating effects of secondary brain insult. For over hundred years, decompressive craniectomy has been a standard surgical intervention for treatment of TBIsowever it is not without harm since it causes serious complications including meningitis, subdural hygroma, hydrocephalous and increased reoperation rate (3,4). Cisternostomy is the most recently introduced intervention for management of cerebral edema. Cisternostomy has proven its efficiency as a standalone reatment and as an adjunctive on decompressive craniectomy in treatment of severe traumatic brain injury. Cisternostomy isperformed by opening and drainage of subarachnoid basal cisterns to atmospheric pressure.This results in opening of the perivascular virchow-Robin Spaces (VRS) thus causing a significant reduction in cerebral edema and hence allows the brain to relax (5–8). This review aims at investigating the therapeutic effects of Cisternostomy when used independently or as adjunctive to decompressive craniectomy (DC) across the available RCTs and Cross sectional studies to optimize the strength of evidence for underpinning the strategy of traumatic brain injury treatment.

### Research questions

1. Does cisternostomy reduce cerebral edema and the associated complications when compared with decompressive craniectomy in managing TBI?
2. Does the cisternostomy influence the length of patient recovery when compared with decompressive craniectomy ?
3. Is there any prognostic difference among patients with severe traumatic brain injury who are managed with cisternostomy versus the arm managed with decompressive craniectomy?

The research questions in this review are designed to be answered according to the PICOS protocol as follows:

### Participants

Human subject, adults including females and males

### Interventions

Cisternostomy, Decompressive Craniectomy DC with Adjunctive Cisternostomy

### Comparators

Cisternostomy versus Decompressive craniectomy

### Outcomes

Recovery time, Length of ICU admission, Ventilation Duration, Length of hospital stay (LOS), Procedural associated complications including cerebral edema meningitis, subdural hygroma, hydrocephalous and reoperation rate.

### Study design

This study is a systematic review with meta-analysis

### Objectives Broad Objective

To evaluate the therapeutic effect of cisternostomy versus decompressive craniectomy in patient with severe TBI

### Specific objectives

1. To evaluate the value of adjunctive cisternostomy in reducing the rate of decompressive craniectomy-related complications
2. To determine the efficacy of cisternostomy and decompressive craniectomy in reducing the intracranial pressure in patients with severe TBI
3. To evaluate the efficiency of cisternostomy and decompressive craniectomy in mitigating the sequelae of secondary TBI
4. To compare the therapeutic outcomes of cisternostomy and decompressive craniectomy in patients with severe TBI

## MATERIALS AND METHODS

### Protocol and Registration

We shall draft and develop the review protocol in accordance to the provisions of the Preferred Reporting Items for Systematic Review and Meta-analysis (PRISMA) 2020 guideline. We will submit the protocol manuscript to PROSPERO for review registration.

### Eligibility Criteria

#### Inclusion Criteria

We shall recruit all primary research articles reporting on the therapeutic effect of cisternostomy, decompressive craniectomy and decompressive craniectomy with adjunctive craniectomy for treatment of traumatic brain injury. We will include both Randomized Clinical Trials (RCTs) and Non Randomized Studies of the effect of Interventions (NRSI) with minimal risk of bias within the range of 10 years publication from January 2013 to December 2022.We will recruit only articles published in English language.

#### Exclusion Criteria

We will exclude animal model-based studies, articles involving the study of pediatric population, articles published in languages other than English, and articles with higher risk of bias (ROB)

### Source of Information

We will perform the literature search from several electronic databases including EMBASE, PubMed, Scopus, Web Of Science (WOS), COCHRAINE, Global Index Medicus, Google Scholar and Semantic Scholar

### Search and Search Strategy Search

Two authors VMK and TMJD will independently perform the literature search from electronic databases using the following key terms in combination with Boolean operators, Cisternostomy [All Fields], Decompressive Craniectomy [MeSH Terms], Decompressive hemicraniectomy [All Fields], Traumatic brain injury[MeSH Terms].

### Search Strategy

Cisternostomy [All Fields] AND decompressive craniectomy [MeSH Terms] OR decompressive hemicraniectomy [All Fields] AND traumatic brain injury [MeSH Terms] .The search strategy for PubMed electronic database is annexed **table 1**.

**Table 1:**
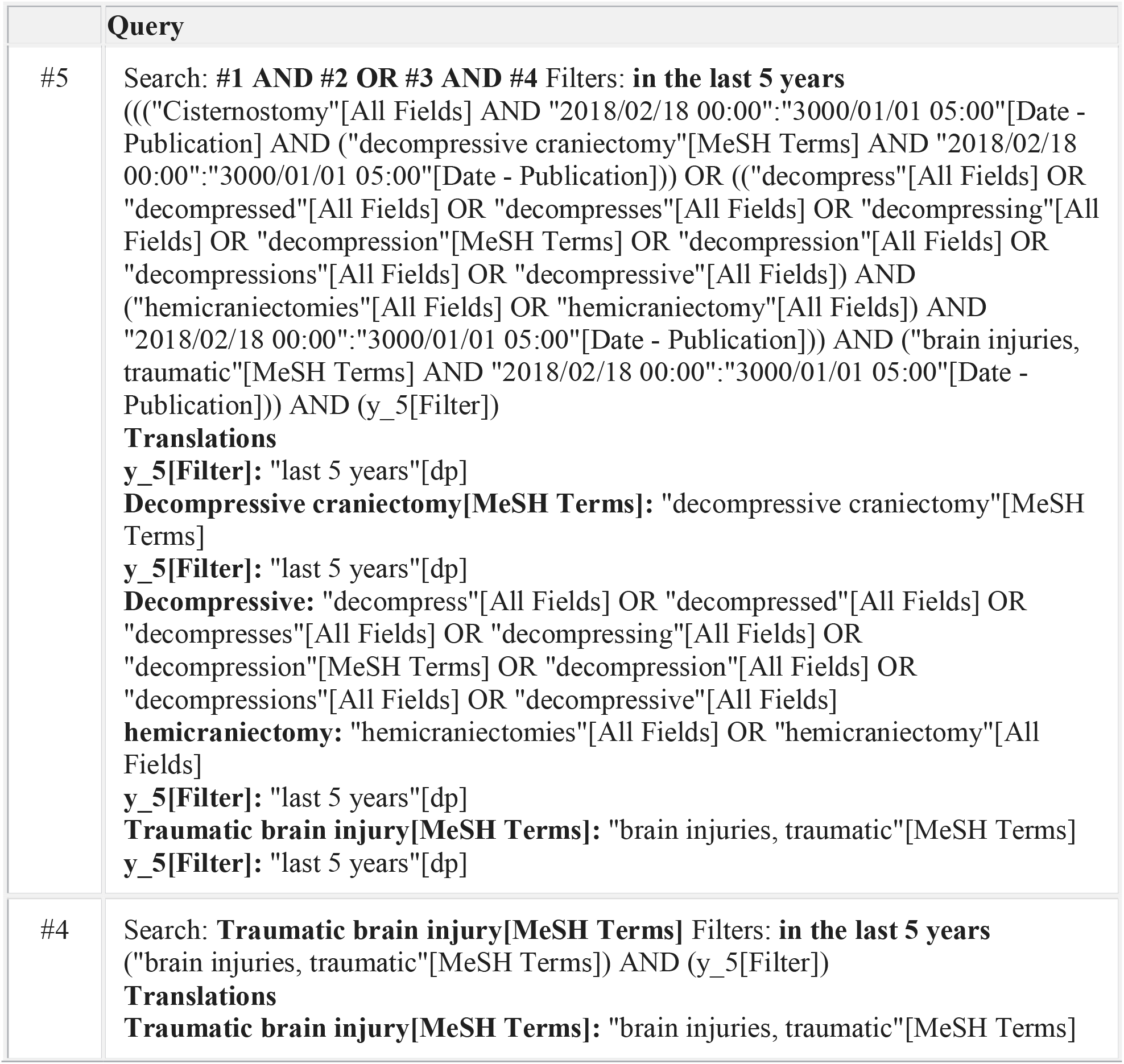
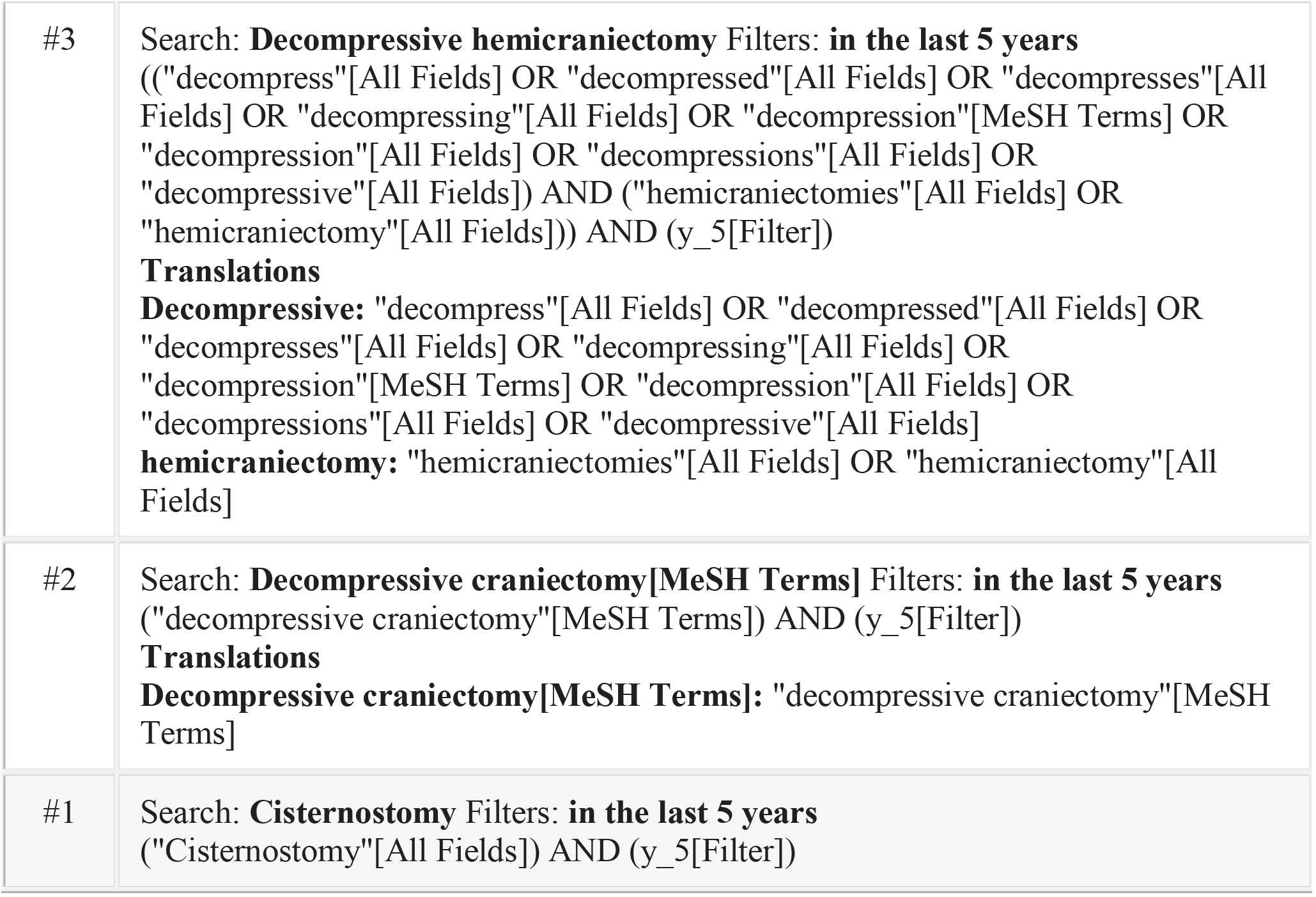
PubMed electronic database search strategy

**Table 2:** Data category items

| S/N | Category of data item | Variable (Item) |
| --- | --- | --- |
| 1. | Baseline characteristics of participants | Age , Sex , TBI severity status |
| 2. | Article content and publication information | Authors , Study design i.e RCT or NRSI , Sample Size , Study period , Year of publication |
| 3. | Intervention | Cisternostomy , Decompressive craniectomy and DC adjunct Cisternostomy |
| 4. | Therapeutic outcomes | Effect on ICP , Cerebral edema , Recovery status , Duration of admission to ICU , Duration on Mechanical ventilation , Complications reported |

### Study Selection

Two reviewers (VMK and TMJD) will independently perform both the primary and full text screening of all articles and the entire process of screening will be blinded. The articles will be selected for review based on the recruitment criteria. Any conflict of interest upon data screening will be resolved by assentment or resolution from a third reviewer (LKK).

We will use he Rayyan or Covidence software to achieve the data screening and the analysis will be performed using the Review Manager (RevMan) Software. The screening results will be summarized on the PRISMA flow diagram; the reasons for article exclusion will be justified.

### Data Extraction

The data will be extracted from full text screened articles, by the reviewers VMK and TMJD, using a pretested Microsoft Excel sheets. All discrepancies encountered in the course of data extraction process will be mitigated by assentment or resolution from the third reviewer LKK.

### Data Items

#### Risk of bias (ROB) in individual studies

Two reviewers (VMK and TMJD) will evaluate using the ROB tool Version 2 for RCT studies (9) and Risk of Bias in Non-randomized Studies of Interventions (ROBINS-I) for NRSI designs and the overall ROB will be rated as low risk, moderate risk, serious risk and critical risk (10).

#### Measurement of effect

We will measure the effect of intervention for therapeutic effect and outcome using the Hazard Ratio (HR) for RCTs and Relative Risk and Propensity Matching Analysis for NRSI. The Mean Difference (MD) and Standard Mean Difference (SMD) for continuous variables will be analyzed to estimate the effect size within the population groups at 95% CI

#### Data Synthesis

The Meta-analysis will be performed for at least more than 5 articles with minimal heterogeneity and low ROB .The data pertaining to effect of cisternostomy, decompressive craniectomy and cisternostomy adjunctive to decompressive craniectomy will be generated and analyzed, the Random Effect Model (REM) of each intervention will be presented.

The consistency of the data from all articles will be evaluated for clinical heterogeneity, methods heterogeneity, statistical heterogeneity and publication bias. The Funnel plot analysis will be performed to evaluate heterogeneity including Chi^2^-test and I^2^.If significant heterogeneity will be detected from the data only qualitative analysis will be conducted with adherence to Synthesis Without Meta-analysis (SWiM) protocol. The Mata-analysis will be performed using Reviw Manager (RevMan) software.

#### Risk of bias across studies

If more than 5 articles will be extracted for Meta-analysis, the ROB across the articles will be analyzed using the funnel plot.

#### Subgroup Analysis

For significant heterogeneity across the data, the specific subgroups including cisternostomy, decompressive craniectomy and cisternostomy adjunctive to decompressive craniectomy, severity of TBI, prognostic status.

#### Sensitivity Analysis

To assess the robustness of synthesized data and confounders, we will perform the sensitivity analysis by omitting the articles with high ROB, controversial definition of effect of interventions and high risk of bias.

#### Strength of body of evidence

The strength and external applicability of the synthesized evidence will be evaluated by two reviewers (VMK and LK) using the Grading of Recommendation, Assessment, Development and Evaluation (GRADE) protocol. The quality levels of evidence will be classified as high, moderate, low and very low level of evidence (LOE) according to the GRADE provisions (11,12).

#### Ethical consideration and Dissemination

This review will not include any human participant such that the ethical clearance approval is not applicable. The protocol of this review will be registered at PROSPERO registry.We will disseminate the final report of this review to local and international scientific conferences. The manuscripts for this protocol and final result will be submitted for publication in the reputable Journal of Neurotrauma.

## Data Availability

Not applicable

## DECLARATION

### Contribution of Authors

Preparation of the full text of this protocol: VMK and TMJD

Concept and Context: VMK, TMJD and LKK

Drafting of Manuscript: VMK, TMJD and LKK

Literature Search: VMK and TMJD

Critical revision of the manuscript for intellectual content: All authors

Supervision: FG, IE and OA

## APPENDIX

